# Discrimination of breast cancer from healthy breast tissues using a three-component diffusion-weighted MRI model

**DOI:** 10.1101/2020.09.03.20179481

**Authors:** Maren M. Sjaastad Andreassen, Ana E. Rodríguez-Soto, Christopher C. Conlin, Igor Vidić, Tyler M. Seibert, Anne M. Wallace, Somaye Zare, Joshua Kuperman, Boya Abudu, Grace S. Ahn, Michael Hahn, Neil P. Jerome, Agnes Østlie, Tone F. Bathen, Haydee Ojeda-Fournier, Pål Erik Goa, Rebecca Rakow-Penner, Anders M. Dale

## Abstract

**Purpose:** Diffusion-weighted magnetic resonance imaging (DW-MRI) is a contrast-free modality that has demonstrated ability to discriminate between pre-defined benign and malignant breast lesions. However, the ability of DW-MRI to discriminate cancer tissue from all other breast tissues in a clinical setting is unknown. Here we explore the ability to distinguish breast cancer from healthy breast tissues using signal contributions from the newly developed three-component multi-b-value DW-MRI model.

**Experimental design:** Pathology-proven breast cancer patients from two datasets (n=81 and n=25) underwent multi-b-value DW-MRI. The three-component signal contributions C_1_ and C_2_ and their product, C_1_C_2_, were compared to the image defined on maximum b-value (DWI_max_), conventional apparent diffusion coefficient (ADC), and apparent diffusion kurtosis (K_app_). Ability to discriminate between cancer and healthy breast tissues was assessed by the false positive rate given sensitivity of 80% (FPR_80_) and receiver operating characteristic (ROC) area under the curve (AUC).

**Results:** Mean FPR_80_ for both datasets was 0.016 (95%CI=0.008–0.024) for C_1_C_2_, 0.136 (95%CI=0.092–0.180) for C_1_, 0.068 (95%CI=0.049–0.087) for C_2_, 0.159 (95%CI=0.114–0.204) for DWI_max_, 0.731 (95%CI=0.692–0.770) for ADC and 0.684 (95%CI=0.660–0.709) for K_app_. Mean ROC AUC for C_1_C_2_ was 0.984 (95%CI=0.977–0.991).

**Conclusions:** The three-component model yields a clinically useful discrimination between cancer and healthy breast tissues, superior to other DW-MRI methods and obliviating pre-defining lesions by radiologists. This novel DW-MRI method may serve as non-contrast alternative to standard-of-care dynamic contrast-enhanced MRI (DCE-MRI); removing the need to administer Gadolinium contrast decreases scan time and any accumulation of Gadolinium in the brain.

## INTRODUCTION

Numerous studies have indicated that early breast cancer detection, with dynamic contrast-enhanced magnetic resonance imaging (DCE-MRI), has higher sensitivity than current screening programs (ultrasound and mammography)^1-5^. However, DCE-MRI has a number of limitations such as conflicting results regarding specificity^2-6^, dependency on expert radiologist readers, and the use of Gadolinium-based contrast agents that are linked to deposition in the brain^7^, additional scan time and additional costs^8^. In contrast, diffusion-weighted MRI (DW-MRI) does not require exogenous contrast and yields quantitative information of tissues microstructure by detecting diffusion of water molecules through application of varying degree of diffusion weighting (b-values).

Various diffusion models have demonstrated comparable ability to DCE-MRI in discriminating between *pre-defined* benign and malignant lesions in small regions of interest (ROIs) in the breast^9-15^. However, diffusion imaging-based tools are underexplored for discrimination of cancer tissue from all benign breast tissue (benign lesions and normal fat and fibroglandular tissue), which would increase its clinical utility and practicality in breast cancer screening, treatment evaluation, surgical planning, and surveillance, where lesions are not pre-defined by radiologists.

Multi-component modeling of the DW-MRI signal over extended ranges of b-values (typically up to 2000-3000 s/mm^2^) theoretically isolates the slowly diffusing water component present in cancer tissue in several organs^16-20^. Each component is defined by an apparent diffusion coefficient (ADC), and therefore represents a distinct pool of the water diffusion signal, with the corresponding magnitude. Vidić et al^13^ demonstrated that the normalized magnitude of the slowest element in a two-component model was excellent (AUC=0.99) in discriminating between benign and malignant breast lesions. Further, when optimizing the multi-component model to fit the DW-

MRI signal from all breast tissues, including cancer and healthy breast tissues, the result was a three-component model with empirical ADCs determined across patients and sites^21^. The main objective of the current study is to explore the ability of estimates derived from a three-component model to discriminate breast cancer from healthy breast tissues and to compare it to conventional DW-MRI imaging.

## PATIENTS AND METHODS

### Patients

In order to validate the discriminatory power of the three-component model across scanners and sites, two datasets of pathology-proven breast cancer patients from a United States (US) site (n=81) and a European site (n=25) were included (Table 1). Note that 49 cases from the US site and all cases from the European site were also used to determine the three-component model with fixed ADCs for breast tissue^21^. In addition, cases from the European site have been previously used for DW-MRI modeling of previously defined benign and malignant lesions^13,22-25^, linking DW-MRI signal to histological specimen^26^ and distortion correction techniques^27^.

**Table 1.**
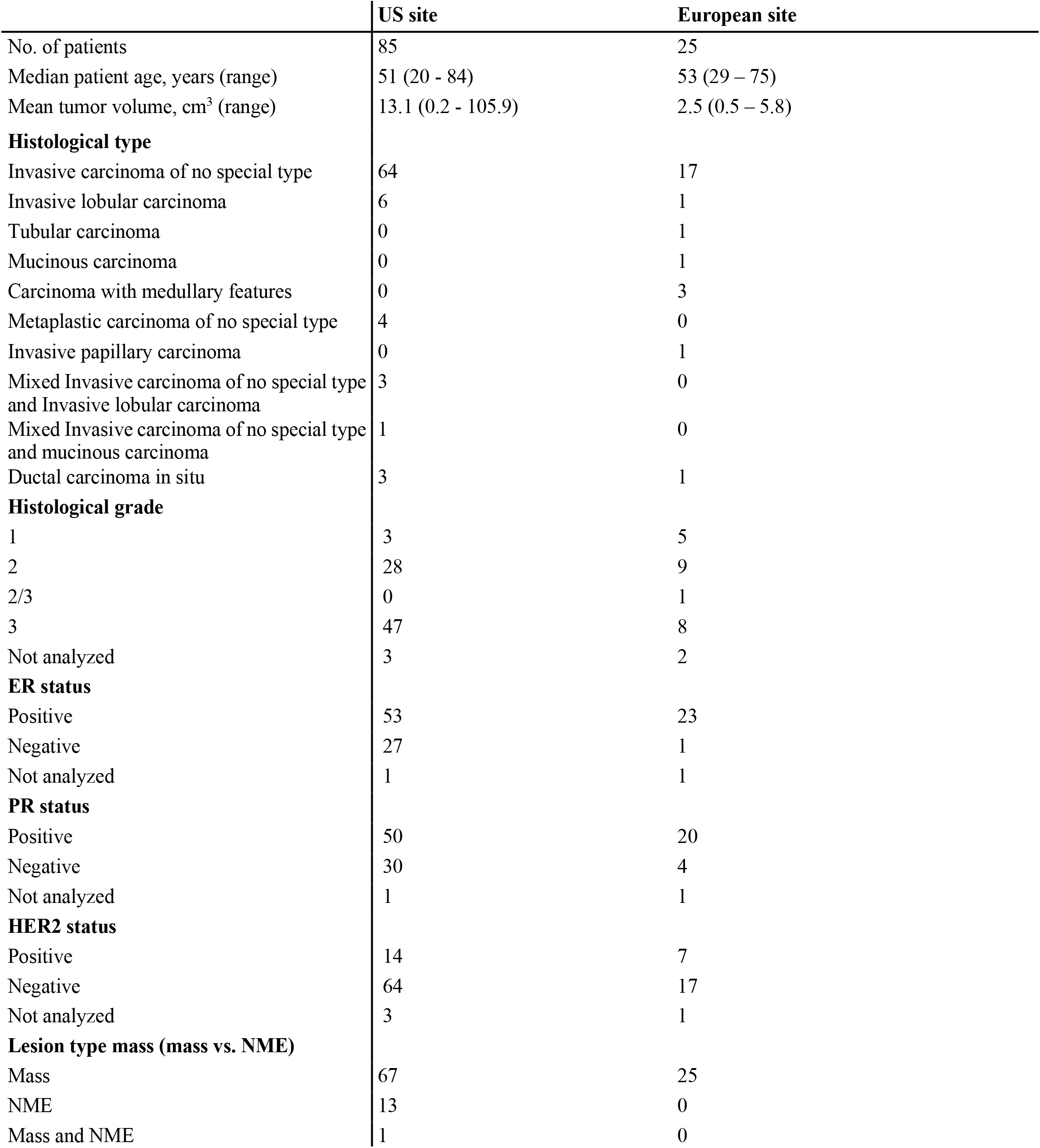
Table of patient characteristics. ER (estrogen receptor) and PR (progesterone receptor) status were assessed by immunohistochemistry (IHC) and was considered positive if ≥1% stained nuclei was present in 10 high-power fields^35^. Human epidermal growth factor receptor 2 (HER2) status was assessed by ICH and fluorescence in situ hybridization (FISH) according to ASCO/CAP guidelines 2013^36^ or 2018^37^ (depending on time of recruitment); positivity was defined as an IHC score of 3+, or 2+ with a gene to chromosome ratio ≥2.0 by FISH. ER, estrogen receptor; HER2, human epidermal growth factor receptor 2; NME, non-mass enhancement; PR, progesterone receptor.

#### US site

Patients with pathology-proven breast cancer with no cytotoxic regimens, chemotherapy, or ipsilateral radiation therapy for this malignancy prior to MRI scanning were eligible for this prospective study. The study was approved by the Institutional Review Board of the US site. The recruitment of patients began in December 2015 and ended in June 2019. Tumor categorization was done by histopathologic analysis of core needle and open incisional biopsies. In total, 14 patients were excluded from the study; nine patients had contralateral cancer or mastectomy, one patient had no visible cancerous tissue on DW-MRI, and in four patients image quality was low (low signal-to-noise ratio (n=2), poor fat saturation (n=1), and severe image distortion (n=1)), resulting in 81 patients.

### European site

This prospective study was approved by the Regional Committees for Medical and Health Research Ethics (REC Central Norway, 2011/568). The recruitment of patients began in August 2014 and ended in August 2016. Inclusion criteria and tumor categorization were similar to those of the US site; for more details, see inclusion of malignant lesions from Vidić et al^13^.

### MRI Acquisition

MRI data were acquired on a 3T GE scanner (MR750, DV25-26, GE Healthcare, Milwaukee, United States) and an 8-channel breast array coil with a bilateral axial imaging plane for the US dataset, while patients from the European dataset were imaged with a 3T Siemens scanner (Skyra, VD13-E11, Siemens Healthcare, Erlangen, Germany) and a 16-channel breast array coil with a unilateral sagittal imaging plane. Differences in scanner and pulse sequence parameters across sites were used to determine that the discriminatory potential of the three-component model is robust for data collected in different scanners and pulse sequence parameters. In addition to Gadolinium DCE-MRI and T_2_-images, both datasets included high b-value DW-MRI acquisition:

#### US site protocol

Bilateral axial DW-MRI was performed using reduced field of view (FOV) echo-planar imaging (EPI) including the following parameters: spectral attenuated inversion recovery (SPAIR) fat suppression, TE=82 ms, TR=9000 ms, b-values (number of diffusion directions)=0, 500 (6), 1500 (6), and 4000 (15) s/mm^2^, F0V=160×320 mm^2^, acquisition matrix=48×96, reconstruction matrix=128×128, voxel size=2.5×2.5×5.0 mm^3^, phase-encoding (PE) direction A/P, and no parallel imaging.

### European site protocol

Unilateral sagittal DW-MRI was performed using Stejskal-Tanner spin-echo EPI including the following parameters: FatSat (n=15) and SPAIR (n=10) fat suppression, TE=88 ms, TR=10,600 ms (n=15) and 11,800 ms (n=10), b-values (number of diffusion directions)=0, 200 (6), 600 (6), 1200 (6), 1800 (6), 2400 (6), and 3000 (6) s/mm^2^, F0V=180×180 mm^2^, acquisition matrix=90×90, reconstruction matrix=90×90, voxel size=2.0×2.0×2.5 mm^3^, PE direction A/P, generalized auto-calibrating partially parallel acquisition (GRAPPA) with acceleration factor of 2 and 24 reference lines.

### Image Processing and Analysis

Data were normalized to the 98th percentile of intensity within the b=0 s/mm^2^ image, indicated by the normalization factor (N). This was done to address different image intensity scaling while simultaneously preserving contribution of T_2_-decay to DW-MRI signal. Noise correction^28^ was performed to account for decreasing signal to noise ratio with increasing b-value. The observed signal (SI_obs_) is the mean signal across diffusion directions from one individual b-value image. Background voxels were selected by manually placing a ROI in an area in the air outside the breast on the highest b-value image, yielding the mean background intensity (SI_bkg_). The corrected signal intensity (SI_corr_) calculated from SI_obs_ and SI_bkg_ is given as:

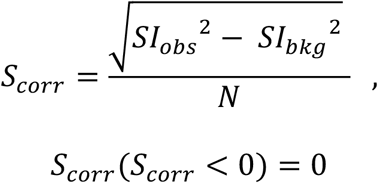

Furthermore, corrections for eddy current artifacts, motion^17^ and geometric distortion^29^ were applied for the European dataset.

Full-volume cancer and control ROIs were manually defined on DW-MRI images, guided by all available data in the exam protocol (including DCE-MRI and anatomical T_2_-images, Figure 1), under supervision of and validation by two breast radiologists: RRP (US site) and AØ (European site). Cancer ROIs were drawn for the lesions corresponding to pathology-proven cancer. Control ROIs were drawn for the entire contralateral breast (US site) and in a cancer-free region in the ipsilateral breast at least 10 mm away from the cancer ROI (European site), with the aim to include all representative healthy breast tissues, excluding the axillary region, large cysts (>2.5 cm), and susceptibility artifacts. Cancer and control ROIs were used to determine discriminatory performance between cancer and healthy breast tissues, respectively.

**Figure 1.**
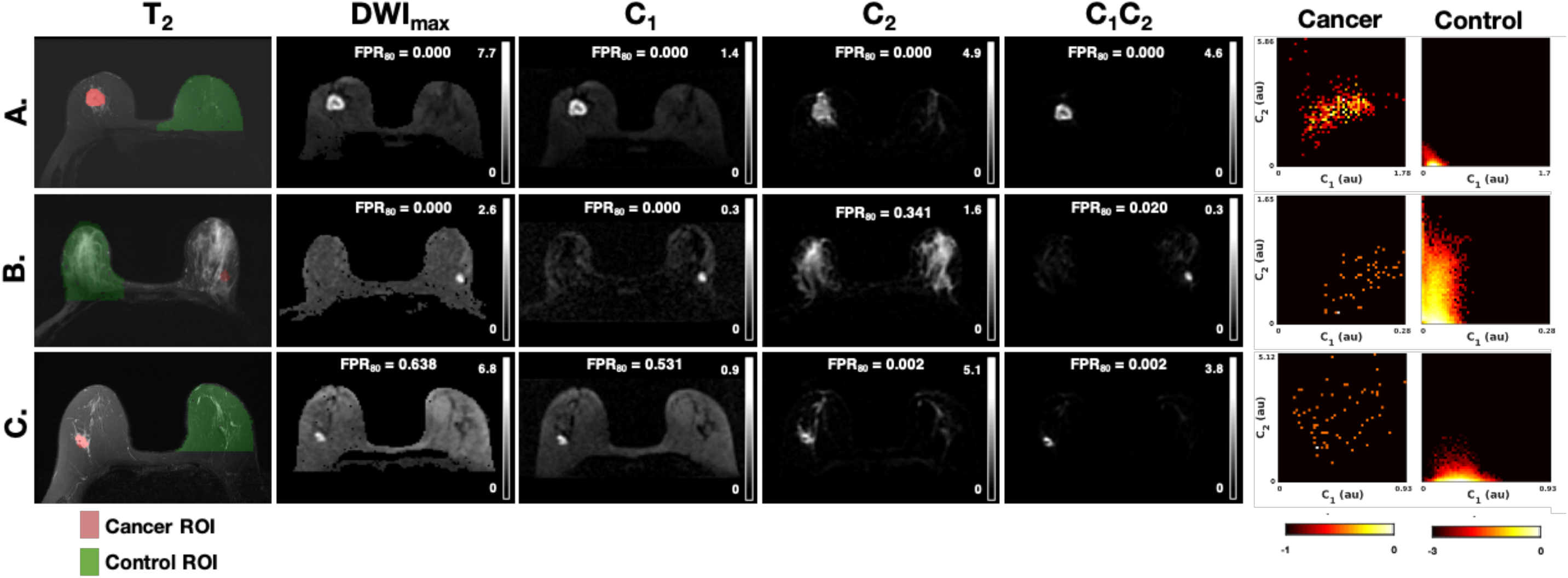
Parameter maps for DWI_max_, C_1_, C_2_, C_1_C_2_ with FPR_80_, T_2_-images with cancer (red) and control (green) ROI overlay and probability density colormaps for cancer and control given C_1_ and C_2_ for three representative cases from the US dataset. FPR_80_ vary depending on healthy tissue composition in relation to the magnitude of C_1_ and C_2_ in cancer tissue. (A.) mixed tissue composition with cancer high on both dimensions. (B.) abundant fibroglandular tissue and high C_1_-magnitude of cancer. (C.) abundant fat tissue and high C_2_-magnitude of cancer. DWI_max_ and C_1_ performance is poorest in (B.), C_2_ in (C.) while C_1_C_2_ has perfect performance across cases. Colormaps are given on a logarithmic scale normalized to the maximum probability density value. Y- and x-axis are defined by the maximum value for each case. Grey level windows for all images are scaled to the maximum and minimum signal intensity of each case and given in arbitrary unites. Au, arbitrary unit; C, signal contribution; DWI_max_, image defined on maximum b-value; FPR_80_, false positive rate given sensitivity 80%; ROI, region-of-interest.

For comparison with other DW-MRI methods, the non-noise-corrected image defined on maximum b-value (DWI_max_), conventional apparent diffusion coefficient (ADC), and apparent diffusion kurtosis (K_app_) were estimated. DWI_max_ was acquired at b=4000 s/mm^2^ for the US dataset and b=3000 s/mm^2^ for the European dataset. The exponential decay formulas described by Jensen et al^30^ and the corresponding b-value limits, <1000 s/mm^2^ and <2000 s/mm^2^, were used for computation of ADC and K_app_, respectively. Note that ADC and K_app_ were normalized by signal at b=0 per voxel to eliminate T_2_-dependence^30^ rather than dividing all voxels by the same normalization factor to preserve T_2_ information as previously described.

To ensure that regions outside of the breast were not included in analysis, control ROIs were masked using intensity thresholding and 3D connected components (US site) or manually delineated within the breast boundary (European site) and reviewed by RRP (US site) and AØ (European site) (Figure 1 and Figure 4). Additionally, all undefined values (zero and infinite) on the image defined on b=0 s/mm^2^, ADC and K_app_ were excluded.

### Three-component modeling of DW-MRI signal

The DW-MRI signal across all available b-values was fit with a tri-exponential model where the slowest exponential had a fixed ADC of zero, yielding a three-component model with two exponentials and one constant term. The three-component model was chosen because it represents the best fit across all voxels from both healthy breast tissues and cancer tissue determined across patients and sites^21^, given as:

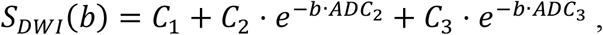

where *S_DWI_* is the diffusion signal in arbitrary units, *b* is the b-value in s/mm^2^, and *C_i_* denotes the voxel-wise unit-less signal contribution of each component. The ADC values were fixed as previously reported in^21^: *ADC_1_*=0 mm^2^/s and therefore not expressed as an exponential, *ADC_2_*=1.4×10^-3^ mm^2^/s, and *ADC_3_*=10.2×10^-9^ mm^2^/s Fixing ADCs ensures comparability of signal contributions across voxels and patients and avoids over-fitting.

The following parametric maps were estimated from the three-component model: C_1_C_2_, C_1_ and C_2_. The parameters C_1_ and C_2_ were estimated directly from the model, while C_1_C_2_ is the corresponding product. The parametric map C_3_ was not included due to low cancer conspicuity^21^.

### Discriminating performance between cancer and healthy breast tissues

Clinical utility of the three-component derived parametric maps was assessed by comparing the discriminatory performance between cancer and healthy breast tissues ROIs of C_1_C_2_, C_1_ and C_2_ maps, DWI_max_, ADC, and K_app_. Because there were ~52 times more healthy breast tissue voxels than cancer voxels, performance in discriminating between cancer and healthy breast tissues was examined for all voxels by the expected false positive rate given a sensitivity of 80% (FPR_80_). In addition, the conventional performance measures receiver operating characteristic (ROC) area under the curve (AUC), sensitivity, specificity, and accuracy were estimated. Sensitivity, specificity, and accuracy were calculated for the threshold value providing optimal accuracy, defined as the mean sensitivity and specificity, assuming equal prevalence of cancer and healthy breast tissue voxels. C_1_C_2_, C_1_, C_2_, DWI_max_, and K_app_^10-12^ were assumed to have higher intensity for cancer compared to healthy breast tissues, while the opposite was assumed for ADC^9^.

## RESULTS

### Sample

The total number of cancer voxels and healthy tissue voxels from both datasets was 37,659 and 1,946,186, respectively.

### Optimized three-component model parameters for discrimination (C_1_, C_2_, and C_1_C_2_)

Probability density colormaps for the three-component model given C_1_ and C_2_ including all voxels across patients and datasets are plotted for cancer (cancer ROIs, Figure 1A) and healthy breast tissues (control ROIs, Figure 1B). These maps display two distinct probability density distributions for cancer and healthy breast tissues. The product C_1_C_2_ discriminates cancer from healthy breast tissues voxels, where voxels low on one or two dimensions corresponds to healthy breast tissues voxels, while cancer probability increases with increased magnitude on C_1_ and C_2_.

The relationship between C_1_ and C_2_ demonstrates that voxels with high magnitude on both dimensions had the highest probability of cancer (Figure 1A); representative cases are given in Figure 1, and all cases are given in Supplementary Figure 1-106. Discrimination performance varied depending on healthy tissue composition in relation to the magnitude of C_1_ and C_2_ in cancer tissue. FPR_80_ was higher (indicating more false positive voxels) for C_1_ and DWI_max_ in a case with abundant fibroglandular tissue and high C_1_-magnitude of corresponding cancer tissue (Figure 1B), compared to abundant suppressed fat tissue and high C_2_-magnitude of corresponding cancer (Figure 1C). The opposite was seen for C_2_, while C_1_C_2_ suppressed both fibroglandular and fatty tissue. This shows that the C_1_C_2_ parameter derived from the three-component model provided the optimal discrimination performance between cancer and healthy breast tissues.

### Discriminatory performance of C_1_C_2_ compared to other DW-MRI methods

Mean FPR_80_ for both datasets was 0.016 (95%CI=0.008–0.024) for C_1_C_2_, 0.136 (95%CI=0.092–0.180) for C_1_, 0.068 (95%CI=0.049–0.087) for C_2_, 0.159 (95%CI=0.114–0.204) for DWI_max_, 0.731 (95%CI=0.692–0.770) for ADC and 0.684 (95%CI=0.660–0.709) for K_app_ (Figure 3). C_1_C_2_ achieved the lowest FPR_80_ with a mean ROC AUC of 0.984 (95%CI=0.977–0.991) when compared to conventional DW-MRI; see Supplementary Table 1 for all conventional performance measures.

C_1_C_2_ had excellent performance compared to ADC and K_app_ in a wide range of representative cases (Figure 4). DWI_max_ performs well in several cases (Figure 4A-B) but underperforms compared to C_1_C_2_, overall (Figure 3) and particularly in a case with abundant fatty tissue (Figure 4C). In addition, C_1_C_2_ visually improves poor DCE-MRI specificity in a case with marked background parenchymal enhancement (Figure 4D). However, C_1_C_2_ underperforms in cases with sparse signal from cancer tissue, such as case of non-mass enhancement (NME) DCIS (Figure 4E). In this case, all DW-MRI derived maps failed to identify tumor tissue compared to healthy breast tissues. Furthermore, there was high diffusion signal from some healthy tissue components such as cysts (Figure 5B), subareolar ducts (Figure 5A), and partial-volume artifact from the interface of fibroglandular and fatty tissue (Figure 4D, Figure 5C). High diffusion signal from cysts and ducts may be defined as non-suspicious with the assistance of T_2_-images (Figure 5A-B).

**Figure 2.**
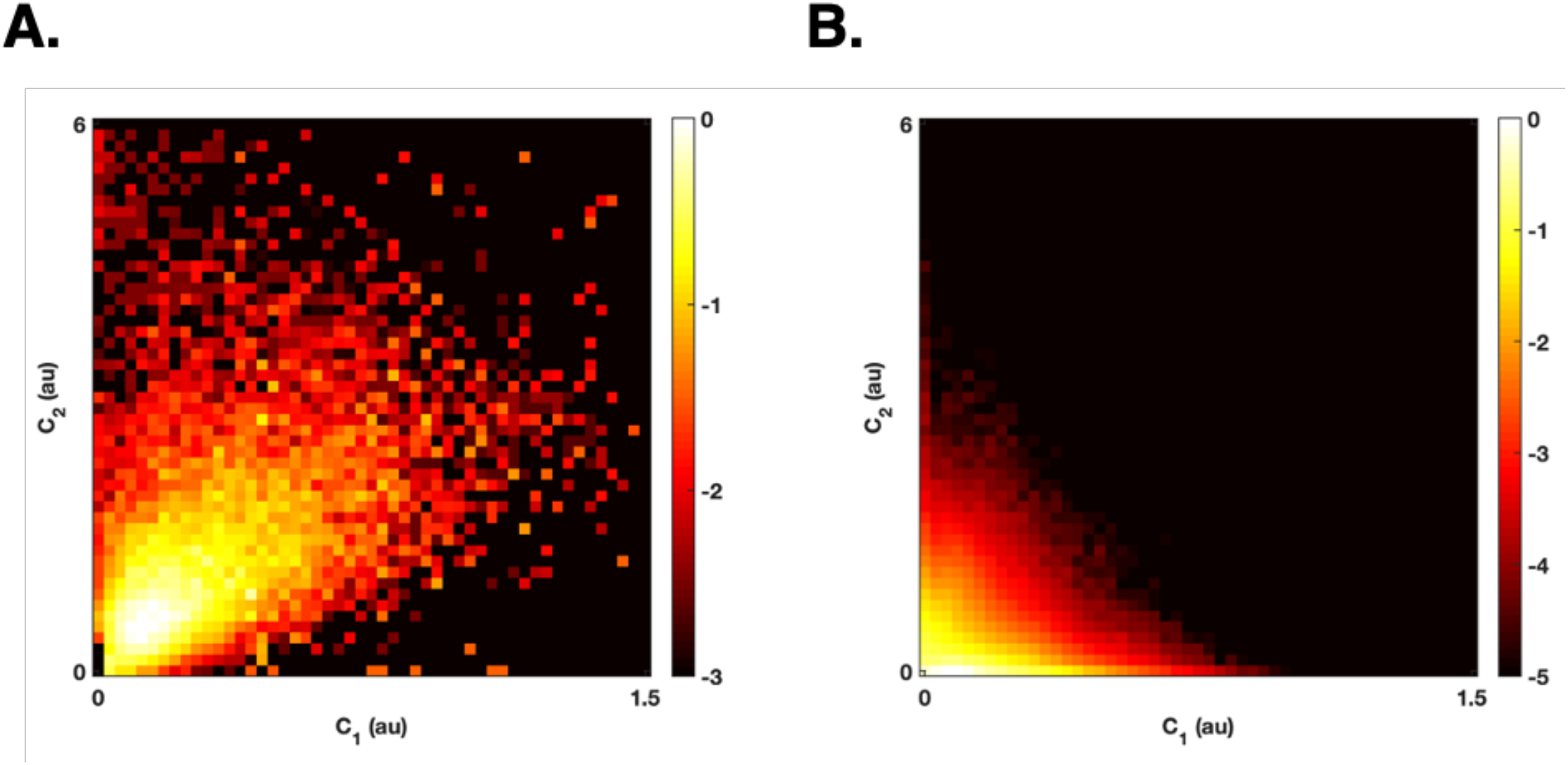
Probability density colormaps for the three-component model given C_1_ and C_2_ including all voxels across patients and datasets are given for (A.) cancer (cancer ROIs) and (B.) healthy breast tissues (control ROIs). These maps display two distinct probability density distributions for cancer and healthy breast tissues. Cancer probability increases with increased magnitude on C_1_ and C_2_. Colormaps are given on a logarithmic scale normalized to the maximum probability density value. Au, arbitrary unit; C, signal contribution.

**Figure 3.**
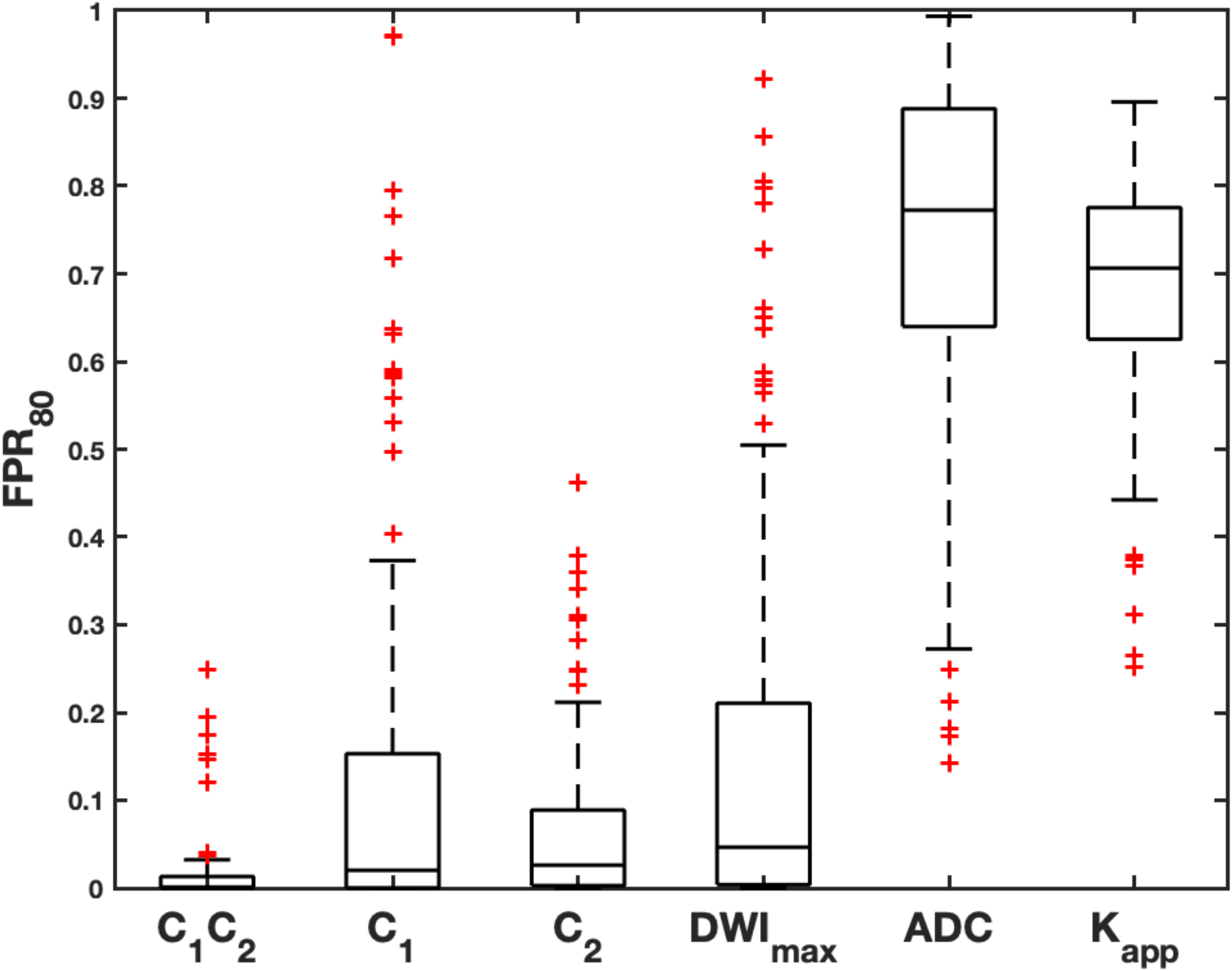
The FPR_80_ is the false positive rate given sensitivity of 80% for discriminating cancer and healthy breast tissues for three-component model parameters (C_1_C_2_, C_1_, C_2_), DWI_max_, ADC and K_app_, given per patient across US and European dataset. Median values indicated by lines; boxes show interquartile range, block bars show data range and red crosses show outliers. The worst FPR_80_ for all maps is 0.9934, which would be 9 934 false positive voxels of one breast (one control ROI) approximated to contain 10 000 voxels (~30 cL). ADC; conventional apparent diffusion coefficient; C, signal contribution; DWI_max_, image defined on maximum b-value; FPR_80_, false positive rate given sensitivity 80%; K_app_, apparent diffusion kurtosis.

**Figure 4.**
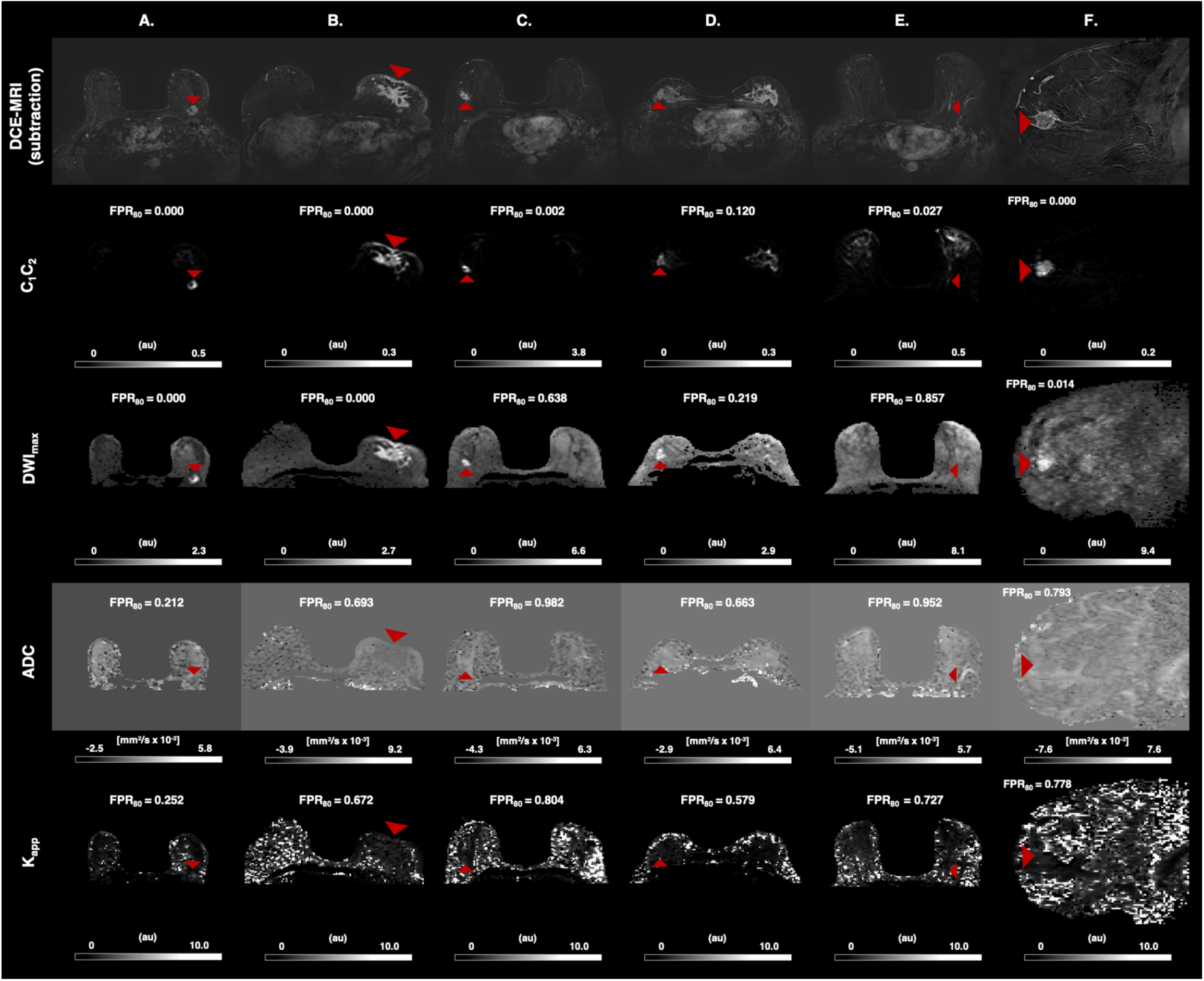
C_1_C_2_, DWI_max_, ADC and K_app_ with FPR_80_ for discrimination between cancer (red arrowhead) and healthy breast tissues (entire cancer-free contralateral breast US dataset, cancer-free ipsilateral breast European dataset) for representative cases from the US (A.-E.) and European (F.) dataset. All cases demonstrate visual similarity between DCE-MRI and C_1_C_2_ maps with excellent performance compared to ADC and K_app_. (A.) Excellent performance by C_1_C_2_ and DWI_max_. (B.) Excellent performance by C_1_C_2 an_d DWI_max_ displaying full extent of cancer involving skin. (C.) Excellent performance by C_1_C_2_ and poor performance by DWI_max_, ADC and K_app_ in a case with abundant fatty tissue. (D.) C_1_C_2_ improves poor DCE-MRI specificity in a case with marked background parenchymal enhancement, but partial-volume artifact from the interface of fibroglandular and fatty tissue in the contralateral breast results in a low discriminatory performance. (E.) A case with NME DCIS where all diffusion maps fail; C_1_C_2_ has reduced cancer signal relative to the high signal from ipsilateral sub-arealor ducts. (F.) Sagittal image plane illustrating same trends in European dataset. The worst FPR_80_ for all maps is 0.9934, which would be 9 934 false positive voxels of one breast (one control ROI) approximated to contain 10 000 voxels (~30 cL). Grey level windows for all images are scaled to the maximum and minimum signal intensity of each case. ADC; conventional apparent diffusion coefficient; Au, arbitrary unit; DCE-MRI, dynamic-contrast enhanced magnetic resonance imaging; DCIS; ductal carcinoma in situ; DWI_max_, image defined on maximum b-value; FPR_80_, false positive rate given sensitivity 80%; K_app_, apparent diffusion kurtosis; NME; non-mass enhancement.

**Figure 5.**
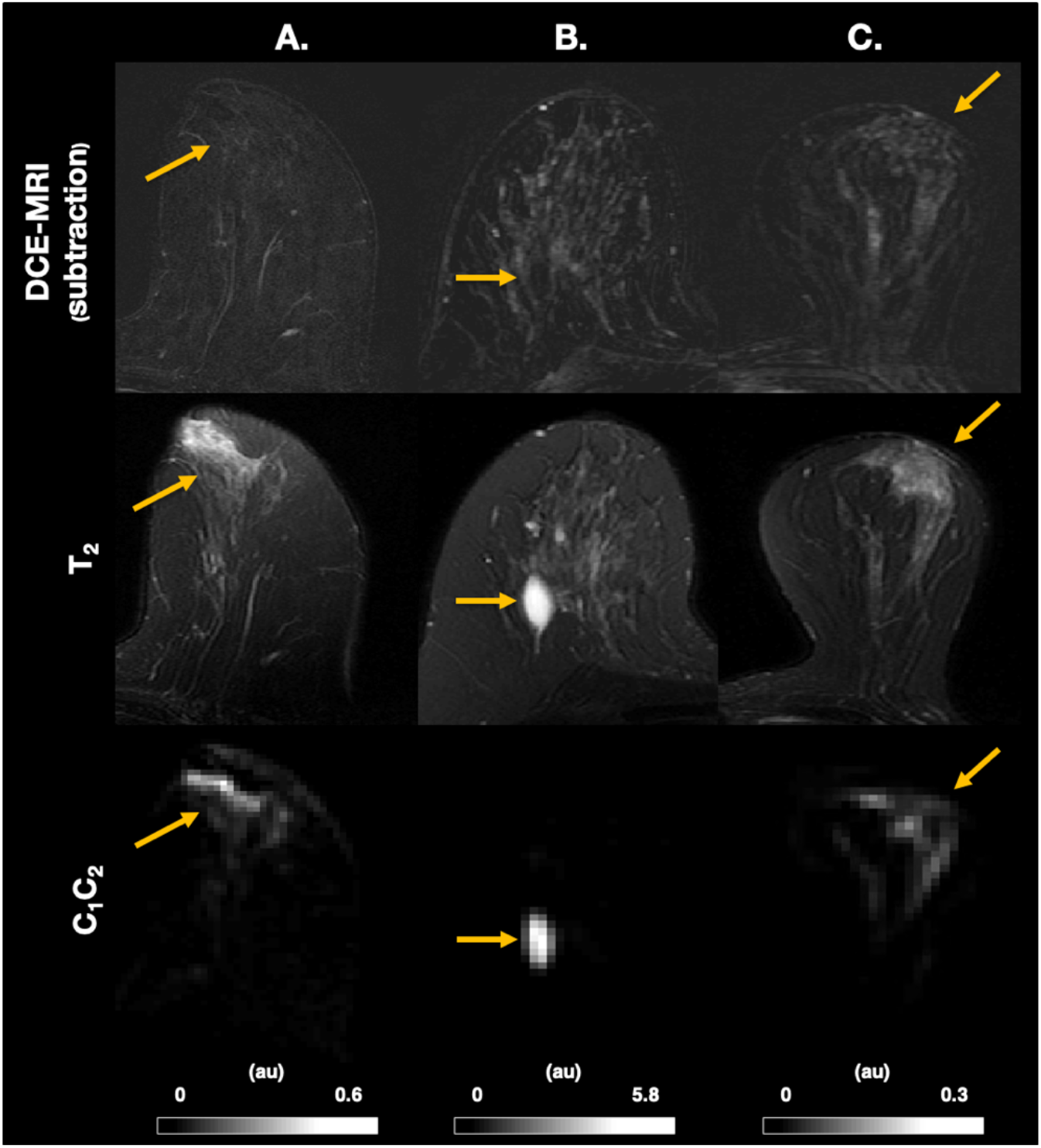
DCE-MRI and T_2_-images with corresponding C_1_C_2_ images illustrating false positives on C_1_C_2_ (yellow arrow). (A.-B.) show that false positive lesions on C_1_C_2_ can be defined as non-suspicious with the assistance of T_2_-images. (A.) High signal involving subareolar ducts on T_2_-image and C_1_C_2_, not visible on DCE-MRI. (B.) Cyst visible on T_2_-image and C_1_C_2_, not visible on DCE-MRI (C.) Demonstration of limitation of C_1_C_2_ where background parenchymal enhancement visible on DCE-MRI and T_2_-image creates a partial-volume artifact corresponding to the interface between fatty and fibroglandular tissue on C_1_C_2_. Grey level windows for all C_1_C_2_ images are scaled to the maximum and minimum signal intensity of each case and given in arbitrary unites. Au, arbitrary unit; C, signal contribution; DCE-MRI, dynamic-contrast enhanced magnetic resonance imaging

## DISCUSSION

Our study shows that breast cancer can be noninvasively discriminated from healthy breast tissues using the derived parameter C_1_C_2_ based on an advanced DW-MRI model, with results comparable to cancer detection using DCE-MRI^2-6^ (FPR_80_ mean=0.016, 95%CI=0.008–0.024 and ROC AUC mean=0.984, 95%CI=0.977–0.991). This means the C_1_C_2_ approach achieved very low false positive rates while detecting 80% or more of the defined cancer voxels. The discriminatory power of the C_1_C_2_ was superior to that of independent signal contributions, conventional DW-MRI-estimates (ADC) and other methods, including kurtosis (K_app_) and DWI_max_. The C_1_C_2_ approach was performed across two different sites, scanners, and acquisition protocols, suggesting potential for real-world applications. The development of this advanced DW-MRI method allows for improved conspicuity of breast cancer relative to background breast tissue. This lays the foundation for a quantitative framework specific to pathology which may serve as an alternative to DCE-MRI.

The high discriminatory performance is due to the novel C_1_C_2_ approach. In addition to malignancy, individual signal contributions from the three-component model were sensitive to the two primary components of healthy breast tissues: fat (C_1_) and fibroglandular (C_2_) tissue. Neither component was sensitive to tissues with very fast diffusion properties, such as vessels, necrosis, or edema. By combining the signal contributions of the two slowest components C_1_ and C_2_, the majority of healthy breast tissues signal was suppressed so that the output image was predominantly sensitive to cancer compared to healthy breast tissues. While optimized for cancer discrimination, the detailed relationship between the three-component model and breast microstructure remains to be studied, as it has been demonstrated for the two-component model^26^.

The C_1_C_2_ approach is performed on data acquired on extended imaging protocols (b-values up to 3000 s/mm^2^ and 4000 s/mm^2^) and requires at least three separate non-zero b-values. Inclusion of higher b-values likely improves discrimination by allowing better estimates of very slow diffusion characteristic of intracellular fluid within hypercellular tumors^13,16-20^. Furthermore, we attribute the high discriminatory performance to the normalization method, where T_2_-contribution to the DW-MRI signal was retained, though noise might also have been a partial contributor. The importance of T_2_ has been demonstrated in separating benign and malignant breast lesions, where the image at b=0 s/mm^2^ yielded a relatively high AUC of 0.85^13^.

We hypothesize that two contributing factors to the poor performance of ADC and K_app_ include the restricted diffusion component from fat tissue and elimination of T_2_-effects that add to cancer discrimination. The FPR_80_ discriminatory performance of ADC and K_app_ varied greatly across subjects; at best, performing around 0.2 in selected cases (Figure 4A), but overall do no better than chance. Conversely, DWI_max_ shares the same basic properties as C_1_C_2_ (diffusion- and T_2_-weighting) and performs noticeably better than ADC and K_app_. However, DWI_max_ is also prone to influence from fatty tissue. The C_1_C_2_ approach better accounts for fatty tissue, conferring a major advantage over DWI_max_ and the other DWI-estimates (Figure 4C), as approximately 50% of women have almost entirely fatty breast tissue or scattered fibroglandular tissue^31^.

In order for C_1_C_2_ to be a noninvasive alternative to DCE-MRI for breast cancer detection, it must have comparable or better sensitivity and specificity. DW-MRI is known to improve detection specificity^9,32^, which is beneficial as lesion-level DCE-MRI specificity have been reported to range from 72-97%^2-6^. In our study, performance was assessed per voxel, and the patient cohort was heterogenous, consisting of a large range of tumor volumes (mean=10.6 cm^3^, range=0.2–105.9 cm^3^), not reflecting the typical patient pool in the screening or surveillance setting which typically have smaller lesions. However, the high performance of discriminating cancer from all other breast tissues in comparison to other DW-MRI-based methods is highly promising and suggests clinical utility comparable to DCE-MRI. C_1_C_2_ may be particularly useful when DCE-MRI demonstrates false positive^33^ and false negative^34^ interpretations in patients with moderate and marked background parenchymal enhancement (Figure 4D). Ideally, C_1_C_2_ may assist in a non-contrast workflow with T_1_- and/or T_2_-sequences as an alternative to DCE-MRI, where false positive findings can be defined as non-suspicious with the assistance of T_2_-images (Figure 5A-B). Thus, successful analysis using C_1_C_2_ removes the need to administer Gadolinium contrast, decreasing scan time and any accumulation of Gadolinium in the brain^7^.

Several diffusion methods aim to isolate the signal from the slowly diffusing water component from cancer tissue by utilizing broad b-value ranges ^10-13,16-20^. Kurtosis imaging is based on a simple mathematical representation of diffusion data where the derived parameter K_app_ has proven potential utility in the breast^10-12^. More advanced, multi-component models with fixed ADCs have been developed to further probe the microstructure in the brain and prostate: restriction spectrum imaging (RSI, on which the C_1_C_2_ approach is based)^16,17^, the VERDICT model^19^, and the hybrid multidimensional MRI model^20^. A key difference between RSI/C_1_C_2_ and the hybrid multidimensional MRI model is that the hybrid model does not use global ADCs, making comparison across patients difficult; however, the hybrid model does incorporate multi-echo information not available in our study. Moreover, the T_2_-effects seen in RSI/C_1_C_2_ are removed from the other two modeling approaches, potentially reducing cancer conspicuity. Although the other multi-compartment models have shown promising results as cancer biomarkers in the prostate, for example, these results may be limited in breast, where fatty tissue is an important component of healthy breast tissues.

There were some limitations to our study. First, the C_1_C_2_ methodology did not correct for partial-volume artifacts which occurred at the interface between fatty and fibroglandular tissue (Figure 4D, Figure 5C). Such artifacts have the potential to be corrected, which was not investigated in this study but is an area of interest for future improvement. Another limitation concerned the definition of control regions; although we ensured that all control regions were verified as cancer-free, based on MRI review by an expert breast radiologist (both datasets) and exclusion of cases with pathology-proven contralateral cancer in the US dataset, we cannot know if occult cancer may have been included in the control region. The unilateral European dataset may have been particularly prone to this, as the control ROIs were defined in the same breast as the cancer (this also made the size of control regions dependent on the extent of cancer and thus variable from case to case in that dataset). Lastly, detection performance is commonly evaluated at the lesion level. This study used a voxel-wise false positive rate, FPR_80_, as its performance measure, which does not give an absolute measure comparable to other literature. However, we argue that such a measure is useful from a radiologist’s perspective, because it mimics a breast cancer examination where all voxels in the entire image are used.

In conclusion, our study is the first to demonstrate that the derived parameter C_1_C_2_, which is the product of the two slowest components of a three-component DW-MRI model, yields a clinically useful, noninvasive method for discrimination between cancer and healthy breast tissues.

The model eliminates the need for pre-defined lesions that conventional quantitative DW-MRI metrics use. Together with anatomical images such as T_2_, C_1_C_2_ may assist in a combined, non-contrast workflow which could serve as an alternative to DCE-MRI. The highly promising diagnostic properties were generalized across sites, scanners, and acquisition protocols, which is important for feasibility of large-scale studies for validation in routine breast cancer detection and follow-up in comparison to DCE-MRI.

## Data Availability

we do not have IRB approval to share raw data with general public.

## Acknowledgements/Financial support

California Breast Cancer Research Program Early Career Award, GE Healthcare, Fulbright Scholarship Program, Mid-Norway Regional Health Authority #46084400, University of California Breast Cancer Screening Program #25IB-0056, NIH/NIBIB #K08EB026503.

## Published work

Part of the study has been accepted for presentation at ISMRM, Virtual Exhibition, 8-13. August 2020 (Andreassen MMS et al, Discrimination of breast cancer from healthy breast tissues using a tri-exponential model)

